# Human Parainfluenza 2 & 4: clinical and genetic epidemiology in the UK, 2013-2017, reveals distinct disease features and co-circulating genomic subtypes

**DOI:** 10.1101/2022.01.20.22269219

**Authors:** Akhil Chellapuri, Matthew Smitheman, Joseph G. Chappell, Gemma Clark, Hannah C. Howson-Wells, Louise Berry, Jonathan K. Ball, William L. Irving, Alexander W. Tarr, C. Patrick McClure

**Author notes:** Corresponding author **Address for correspondence:** C. Patrick McClure, University of Nottingham, W/A1328 West Block, A Floor, Queens Medical Centre, Nottingham, NG7 2UH, UK. These authors contributed equally to this work (Akhil Chellapuri, Matthew Smitheman).

## Abstract

Human Parainfluenza viruses (HPIV) are constituted by four members of the genetically distinct genera of Respirovirus (type 1 and 3) and Orthorubulavirus (type 2 and 4), causing significant upper and lower respiratory tract infections in both children and adults worldwide. However, despite frequent molecular diagnosis, they are frequently considered collectively or with HPIV4 overlooked entirely. We therefore investigated clinical and viral epidemiological distinctions of the relatively less prevalent Orthorubulaviruses HPIV2 & 4 at a regional UK hospital across four winter epidemic seasons. HPIV2 & 4 infection was observed across all age groups, but predominantly in children under 9 and adults over 40, with almost twice as many HPIV4 as HPIV2 cases. Fever, abnormal haematology, elevated C-reactive protein and hospital admission were more frequently seen in HPIV2 than HPIV4 infection. Each of the four seasonal peaks of either HPIV2, HPIV4 or both, closely matched that of RSV, occurring in November and December and preceding that of Influenza A. A subset of viruses were partially sequenced, indicating co-circulation of multiple subtypes of both HPIV2 & 4, but with little variation between each epidemic season or from limited global reference sequences.

## Introduction

Human parainfluenza viruses types 1 to 4 (HPIV1-4) are collectively the second most common cause of hospitalisation for children under the age of 5, behind only Respiratory Syncytial Virus (RSV) [1-4]. Symptomatic HPIV infection is observed in both adults and children worldwide, affecting both the upper and lower respiratory tract [5, 6] with varying severity in the immunocompromised and elderly [7, 8]. HPIV2 presents generally with common cold-like symptoms and is a frequent cause of croup in infants [2, 9]. HPIV4 is less well characterised but has been associated with bronchiolitis and pneumonia [5, 10]. Parainfluenza virus infections place a significant burden on the global healthcare system. In the US alone, a 12-year retrospective study estimated hospital charges for children under the age of 5 annually totalled in excess of $42 and $57 million for HPIV-associated bronchiolitis and pneumonia, respectively, with a gross yearly HPIV associated US hospitalisation burden estimated at 62,000 days [11].

HPIVs belong to the single-stranded negative sense RNA *Paramyxoviridae* family and are sub-divided to the *Respirovirus* genus (HPIV1 and 3) of the *Orthoparamyxovirinae* subfamily and the significantly genetically distinct *Orthorubulavirus* genus (HPIV2 and 4) of the *Rubulavirinae* subfamily [12]. HPIV2 and 4 present an orthodox six gene Paramyxovirus genome [12], with particularly the Hemagglutinin-Neuraminidase (HN) and also Fusion (F) envelope genes known to display a higher level of antigenic variation than the structural components of the orthorubulavirus genome, making them a more appropriate focus for epidemiological studies [13-16].

Two nearly identical archetypal strains of HPIV2 were described: Greer in the mid-1950s in the US [17] and Toshiba in the late 1970s in Japan [18]. More recently, additional lineage defining strains Vanderbilt 94 and 98 have been characterised, with maximal dissimilarity rates circa 5% at the amino acid level [13]. In contrast HPIV4 is categorised into two different subtypes (4a and 4b) based on antigenic properties [19, 20], despite presenting apparently less divergent genomes [16] which do not meet criteria for demarcation as separate species [12]. To date no distinction in clinical outcome has been made amongst circulating HPIV2 or 4 strains, further illustrated by phylogenetic studies indicating genetically related clades often contain strains from different seasons and distant geographical origins [15, 16].

Formative viral diagnostic protocols reliant on cell culture (commonly with primary rhesus monkey kidney cells) resulted in low recovery rates, little cytopathic effect and a weak haemadsorption pattern [9, 19, 21]. This has in part led to HPIV4’s frequent omission from standard diagnostic respiratory investigation and derivation of minimal reference genome data, in turn contributing to a reduced comprehension of its epidemiological significance [9, 10, 15, 22]. Higher sensitivity and specificity of reverse transcription PCR (RT-PCR) [23] and its cost-effective ability to rapidly diagnose viral respiratory infection has improved HPIV surveillance in the current millennium. Recent advances in sequencing technologies should further redress this shortfall in genomic reference material [24].

To further increase our knowledge of HPIV2 & 4 clinical epidemiology, and the differences between the two infections, we undertook a retrospective analysis of all RT-PCR positive samples at a regional UK diagnostic laboratory between September 2013 and April 2017. We additionally sequenced a sub-sample of archived genomic extracts to characterise the underlying complementary genetic epidemiology occurring in this study period.

## Methods

### Clinical Specimens and audit

Clinical specimens were obtained from sputum, nasopharyngeal aspirate or throat swab samples from patients with suspected respiratory virus infections in primary and secondary care units in the Nottingham University Hospitals Trust catchment area between 1^st^ September 2013 and 12^th^ April 2017. Nucleic acids were extracted and screened by an in house respiratory virus diagnostic panel and stored as previously described [25] until March 2016 when a commercial screening panel was adopted (Ausdiagnostics), adding Coronavirus and Enterovirus detection [26]. Clinical and demographic information for all HPIV 2 & 4 positive samples was retrieved from the Nottinghamshire Information System database. Use of residual diagnostic nucleic acids and associated anonymized patient information was covered by ethical approval granted to the Nottingham Health Science Biobank Research Tissue Bank, by the North West - Greater Manchester Central Research Ethics Committee, UK, reference 15/NW/0685. All data analyses were performed using Microsoft Excel and GraphPad PRISM 7.04.

### RT-PCR, Primer design and sequencing

Available higher titre HPIV 2 & 4 nucleic acids with a cT value of lower than 33 (HPIV2) or 30 (HPIV4) as determined by in house qRT-PCR or with a value greater than 100 (HPIV2) or 1000 (HPIV4) copies per 10μl RNA, as determined by the AusDiagnostics assay were selected for further investigation. cDNA was synthesized with RNA to cDNA EcoDry™ Premix containing random hexamers (Takara Clontech) as per the manufacturer’s instruction.

1 μl of this RT reaction was used for PCR comprising of 1x HotStarTaq PCR buffer (QIAGEN), 3 pmol each primer, 400 μM total dNTPs, 0.375 U HotStarTaq DNA polymerase (QIAGEN) and molecular grade water in a 15 μl volume, then thermocycled at 95°C for (15 min), followed by 55 cycles at 95°C for 20 sec, annealing temperature (see supplementary Table 2) for 20 sec and 72°C for 60 sec, then a final additional 72°C for 2 min. cDNA integrity was initially assessed with a modified pan-orthorubulavirus assay generating a 224 bp product using primers AVU-RUB-F2 & AVU-RUB-R [27]. All available complete HPIV2 & 4 F & HN gene sequences were downloaded from GenBank in October 2017 (NCBI:txid 1979160 & 1979161 respectively) and used to design species-specific primers targeting the envelope genes. Ultimately, three primer combinations were used to generate amplicons (and the resulting sequence data presented herein): HPIV2_Fs and HPIV2_Fas, HPIV2_Fs2 and HPIV2_Fas2, and HPIV4_Fs and HPIV4_FasINT (Supplementary Table 2). PCR products of expected size when examined by agarose gel electrophoresis were subjected to Sanger sequencing (Source BioScience, Nottingham, UK). Sequence identity was confirmed using NCBI Standard Nucleotide BLAST (BLASTn). Sequences were deposited in GenBank under accession numbers MZ576382:MZ576401 and MZ576402:MZ576430 for HPIV2 and HPIV4 respectively.

### Phylogenetic analysis

Sequences were analysed using MEGA software. Study sequences were aligned using ClustalW with reference sequences retrieved from NCBI databases in June 2021, by BLASTN searching for related sequences with complete coverage across the regions amplified. A test of phylogenetic models indicated the Hasegawa-Kishino-Yano model with gamma distribution best fit HPIV2 and with invariant sites best fit HPIV4 data respectively. Maximum likelihood phylogenetic trees with 1000 bootstraps were constructed using the selected models.

### Statistical analyses

Comparison of clinical parameters was performed using Graphpad Prism (v9.3.1) statistical software. Similar to previous studies [9, 22], binary logistic regression comparing HPIV2 and HPIV4 infections was not conducted due to the occurrence of incomplete data for some individuals, limiting our ability to model the contributions of some parameters to a logistic regression. As such, categorical datasets were compared using either Fisher’s exact test or χ2 tests. Median values of continuous datasets were compared using Mann-Whitney tests.

## Results

### Routine diagnostic surveillance

Within the study period from 1^st^ September 2013 to 11^th^ April 2017, 26,593 unique respiratory specimens were investigated by routine RT-PCR viral screening, of which 38.15 % (10283) in total were positive for one or more viruses (data not shown). Of the positive specimens, 121 (1.18 %) detected HPIV2 and 238 (2.31 %) HPIV4. Additional positive specimens included 30.76% Rhino & enterovirus, 20.16% Respiratory Syncytial Virus, 12.19% Influenza A, 10.23% Adenovirus, 6.23% Human Parainfluenza Type 3, 5.55% Human Metapneumovirus, 5.65% Coronavirus (tested only from March 2016 onwards), 3.28% Influenza B and 2.75% Human Parainfluenza type 1 (data not shown). Deduplication of multi-sampled patients yielded 112 and 199 individuals infected with HPIV2 and HPIV4 respectively. Of these, 41.96% of HPIV2 and 43.72% of HPIV4 individuals were co-infected with another pathogen.

### Seasonality

HPIV2 infections peaked around December in both 2013 and 2015, with approximately 4-fold more cases in 2015 than 2013 (Figure 1). HPIV4 presented a more complex pattern, with significant spikes of cases around December of both 2014 and 2016, but also a smaller increase of positivity was observed in the winter of 2013, comparable in magnitude to the HPIV2 caseload (Figure 1). The epidemic Human orthorubulavirus season generally began in October and ended in January, peaking in November and December of each year, although sporadic cases of both HPIV2 & 4 were detected between the biennial peaks each year (Figure 1).

**Figure 1:**
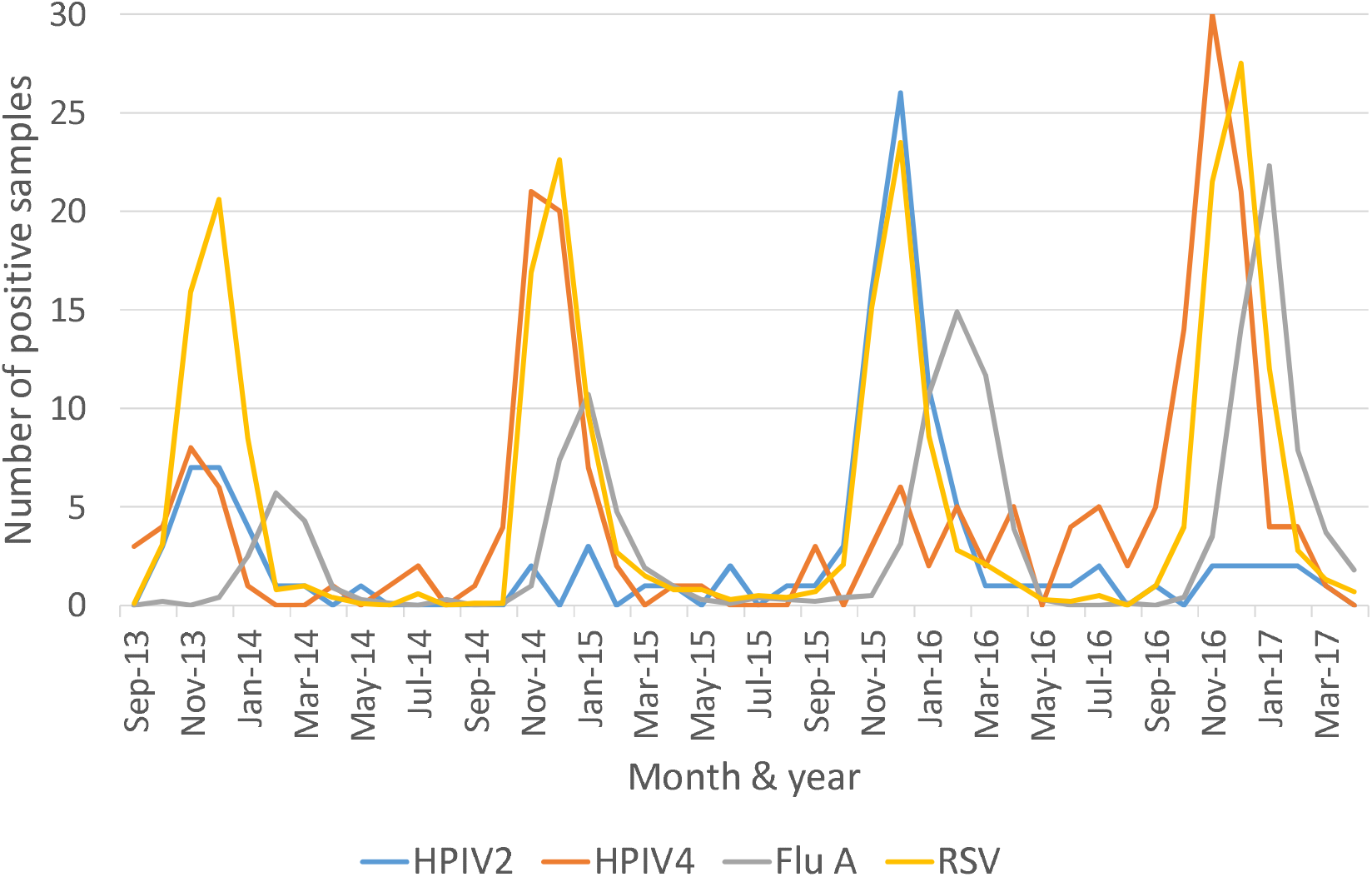
RT-PCR positive HPIV2 & 4 samples compared to Influenza A and RSV by month, between 1^st^ September 2013 and 11^th^ April 2017 in Nottingham UK. RSV and Influenza A numbers have been reduced tenfold to aid clarity.

When compared with the most prevalent seasonal viral pathogens, RSV and Influenza A, each peak of orthorubulavirus cases closely matched those of RSV, although the surge in HPIV4 cases slightly preceded the RSV epidemic in the 2016/17 epidemic period. Both HPIV2 & 4 case spikes always preceded the Influenza A epidemic observed in January and February of each year (Figure 1). Human orthorubulavirus infection appeared independent of co-presentation with RSV, with only 11.6% of HPIV2 and 14.57% of HPIV4 instances of co-positivity (data not shown).

### Demographics

Patients were categorised into gender and six age groups, then assessed for HPIV2 & 4 prevalence (Figure 2). For both types of orthorubulavirus, an increase in cases was observed towards the extremes of age, i.e. in infants / young children and older adults, with low numbers of infections observed in those aged 10 to 40 (adolescents and young adults). This effect was most pronounced in HPIV4 with 61.31% of cases in children under 10 years of age, in contrast to HPIV2 with just 42.86% (Figure 2 and Table 1). The 65 and over elderly group comprised almost one fifth of cases for both HPIV2 (19.64%) and HPIV4 (17.59%, Figure 2 and Table 1).

**Table 1:** Characteristics of hospitalised HPIV2 & HPIV4 mono-infected individuals

|  | **HPIV2** | **HPIV4** | **P Value** |
| --- | --- | --- | --- |
| **Characteristic** | **No. (%)** | **No. (%)** |  |
| **Hospitalised fraction of mono-infected individuals** | **56 (86.1)** | **75 (67.0)** | n/a |
| **Female sex** | 28 (50.0) | 35 (46.7) | 0.7269 |
| **Age (median)** | 21.5 years | 4 | 0.0527 |
| 0-9 years | 24 (42.9) | 47 (62.7) | n/a |
| 10-64 years | 15 (26.8) | 13 (17.3) | n/a |
| 65+ years | 17 (30.4) | 16 (21.3) | n/a |
| **Duration of stay (median)** | 5 days | 5 days | 0.8761 |
| range (days) | 1 - 47 | 1 - 77 | n/a |
| **Underlying medical conditions\*** | 45 (81.82) | 62 (84.93) | 0.6393 |
| **Immunocompromised\*** | 25 (44.64) | 31 (64.58) | 0.0501 |
| **Symptoms** |  |  |  |
| Shortness of breath* | 17 (31.48) | 32 (42.67) | 0.2053 |
| Cough* | 15 (27.78) | 20 (26.67) | >0.9999 |
| Coryza* | 10 (18.52) | 6 (8) | 0.1037 |
| Wheeze* | 1 (1.85) | 6 (8) | 0.2376 |
| Fever* | 26 (48.15) | 11 (14.67) | **<0.0001** |
| Poor feeding* | 3 (5.56) | 6 (8) | 0.7336 |
| **Medical interventions** |  |  |  |
| antibiotics | 25 (48.08) | 42 (57.53) | 0.3636 |
| nebuliser | 5 (9.43) | 9 (12.33) | 0.7761 |
| oxygen | 5 (9.43) | 9 (12.33) | 0.7761 |

*Patients identified between 1<sup>st</sup> September 2013 and 11<sup>th</sup> April 2017. \* Where data* *available for variable; n/a = not applicable; p values <0.05 highlighted in bold*

**Figure 2:**
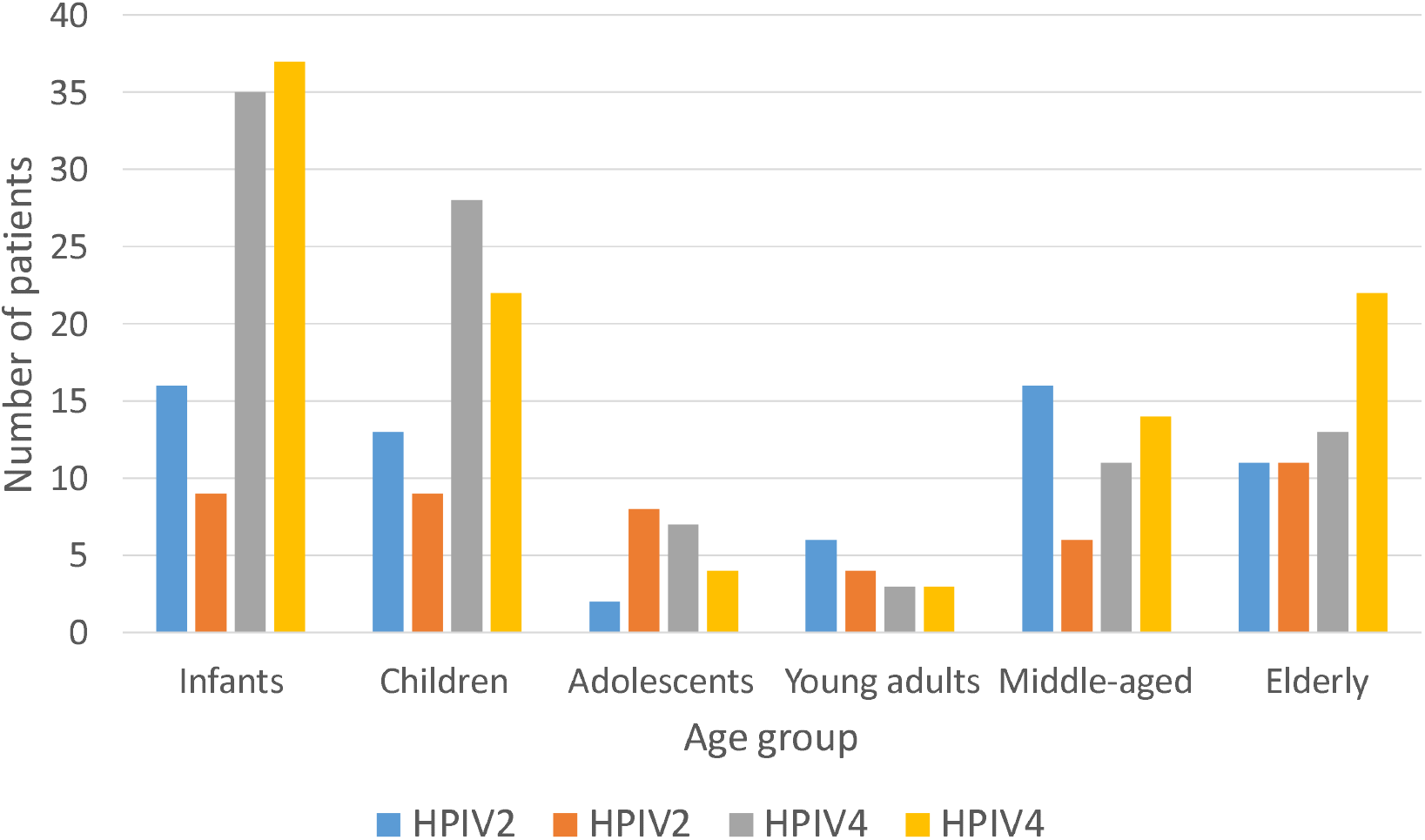
Age and sex distribution of HPIV 2 & 4 positive patients between 1^st^ September 2013 and 11^th^ April 2017. Age group categories are defined by new-born to 1 year old (infants), 2 to 9 (children), 10 to 19 (adolescents), 20 to 40 (young adults), 41 to 64 (Middle aged) and ≥ 65 (elderly).

### Genetic epidemiology

To investigate the underlying genotype of the HPIV2 and 4 positive patients identified, additional RT-PCR was performed on a subsample of higher titre surplus nucleic acid from diagnostic screening and subjected to Sanger sequencing. Available residual samples with higher viral template quantity (see methods) were retrieved and subjected to an array of priming combinations targeting the Fusion (F) and Hemagglutinin (HN) envelope genes, designed with available sequences deposited on GenBank circa September 2017 (data not shown).

Two HPIV2 primer pairs successfully generated overlapping amplicons (1517bp coverage) for the majority of samples tested (20 of 25), allowing phylogenetic analysis of the near complete Fusion gene. HPIV4 amplification proved more challenging, but a single primer pair successfully generated amplicons and 793bp of sequence from the Fusion gene for 29 of 33 samples attempted.

HPIV2 phylogeny in totality was suggestive of three well-supported clades worldwide, defined by the three archetypal strains Greer, V94, V98 [13], but unlike HPIV4 is not formally designated into genotypes (Figure 3). Our recently sampled cohort indicated a predominance of sequences (n = 18 of 20 total) identifying as ‘V94-like’ (red-branched clade, Figure 3), with only two sequences ‘V98-like’ (from patients aged four and 82, pink-branched clade, Figure 3) and none clustering in either the ‘Greer-like’ clade or an additional well-supported clade comprised of two sequences from the USA in 2016 (blue and green-branched clades respectively, Figure 3). Broad distribution across V-94- and V98-like clades was also observed for contemporary HPIV2 reference strains reported predominantly in Seattle, USA and Zagreb, Croatia. However due to availability of residual samples with sufficient viral template, in addition to scarcity of reference HPIV2 strains in general, our data was biased toward the winter 2014/15 season. Only two isolates from outside this period were included (annotated by triangles in Figure 3). A January 2017 sequence was intermingled with majority of the V94-like winter 2014/15 isolates, however an ‘out of season’ HPIV2 from June 2015 appeared genetically distinct from the majority of V94-like isolates, suggesting a further unsampled diversity in HPIV2.

**Figure 3:**
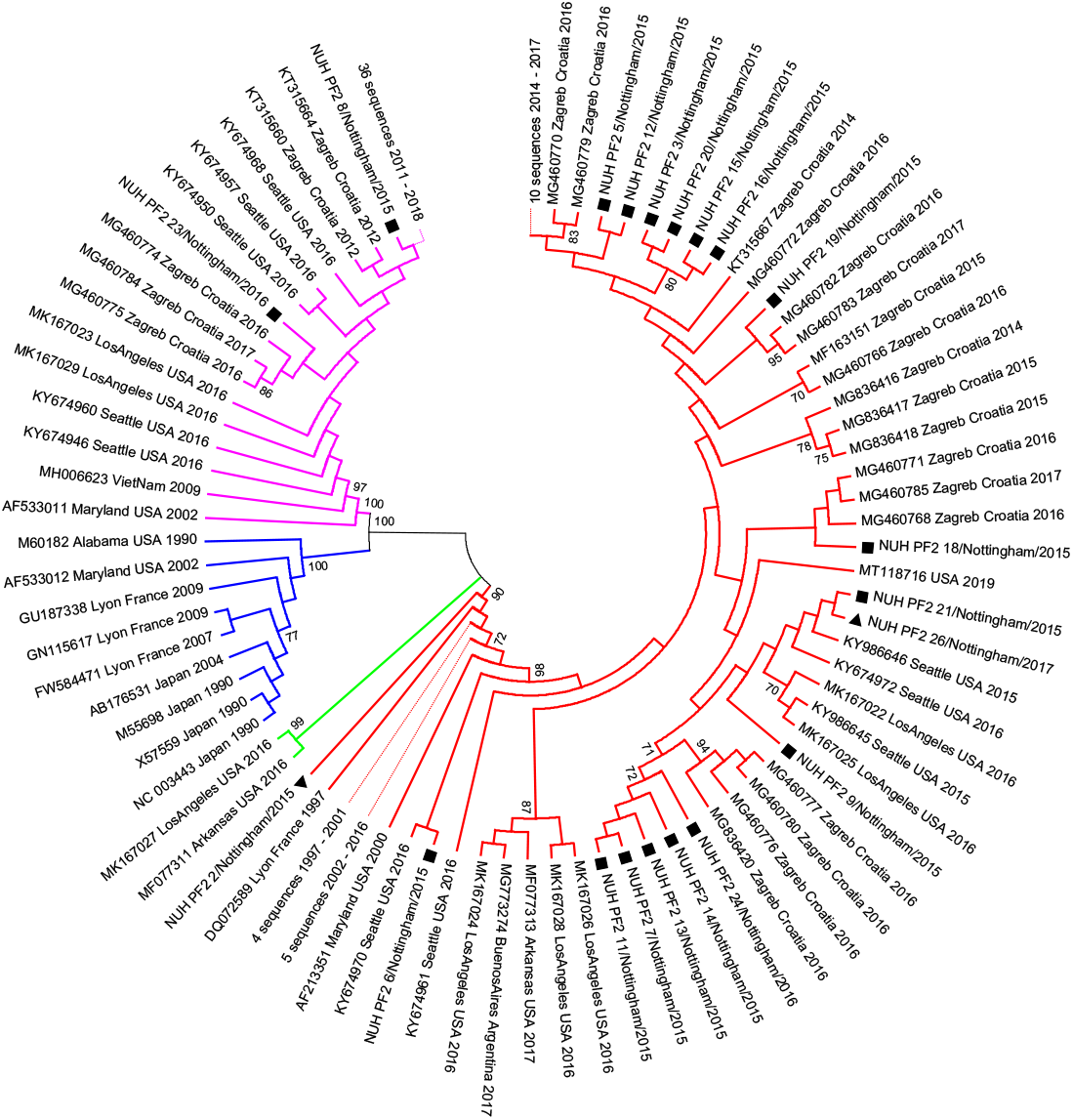
Phylogeny of HPIV2 study strains collected in Nottingham UK between 16^th^ June 2015 and 26^th^ January 2017 compared to available reference sequences derived from GenBank in June 2021 under the taxonomic designation NCBI:txid 2560525. Sequences cover 1517 nucleotides from 4870 to 6386 of reference strain V94, GenBank Accession no. AF533010. The unrooted circular tree was constructed by the maximum likelihood method with 1000 bootstraps, displaying only tree topology; only bootstrap values with support greater than 70 are indicated. Filled squares and triangles represent study samples collected during and outside the core HPIV2 epidemic season respectively. Nominal V98-like (pink), Greer-like (blue), novel (green) and V94-like (red) clade designations are annotated by subtree branch colour; selected subtrees have been collapsed for clarity, with sequence number and year range noted.

Despite both a shorter analysed sequencing region and available reference genomes (28 only as of June 2021) relative to HPIV2, the HPIV4 phylogeny indicated a well-supported division of designated subtypes HPIV4a and b (pink and blue-branched clades respectively, Figure 4). Our sampled isolates from winter 2014 (Figure 4, filled squares) and 2016 (Figure 4, filled triangles) were represented and intermingled in both subtype 4a and 4b. Three ‘out of season’ (Figure 4, open circles) sequences were exclusively 4b, but genetically indistinct from the 2014/15 and 2016/17 winter epidemic season samples. Both HPIV4a and b subtypes appear to further divided by two well-supported distinct lineages, again all represented by sequences derived in different epidemic years (Figure 4).

**Figure 4:**
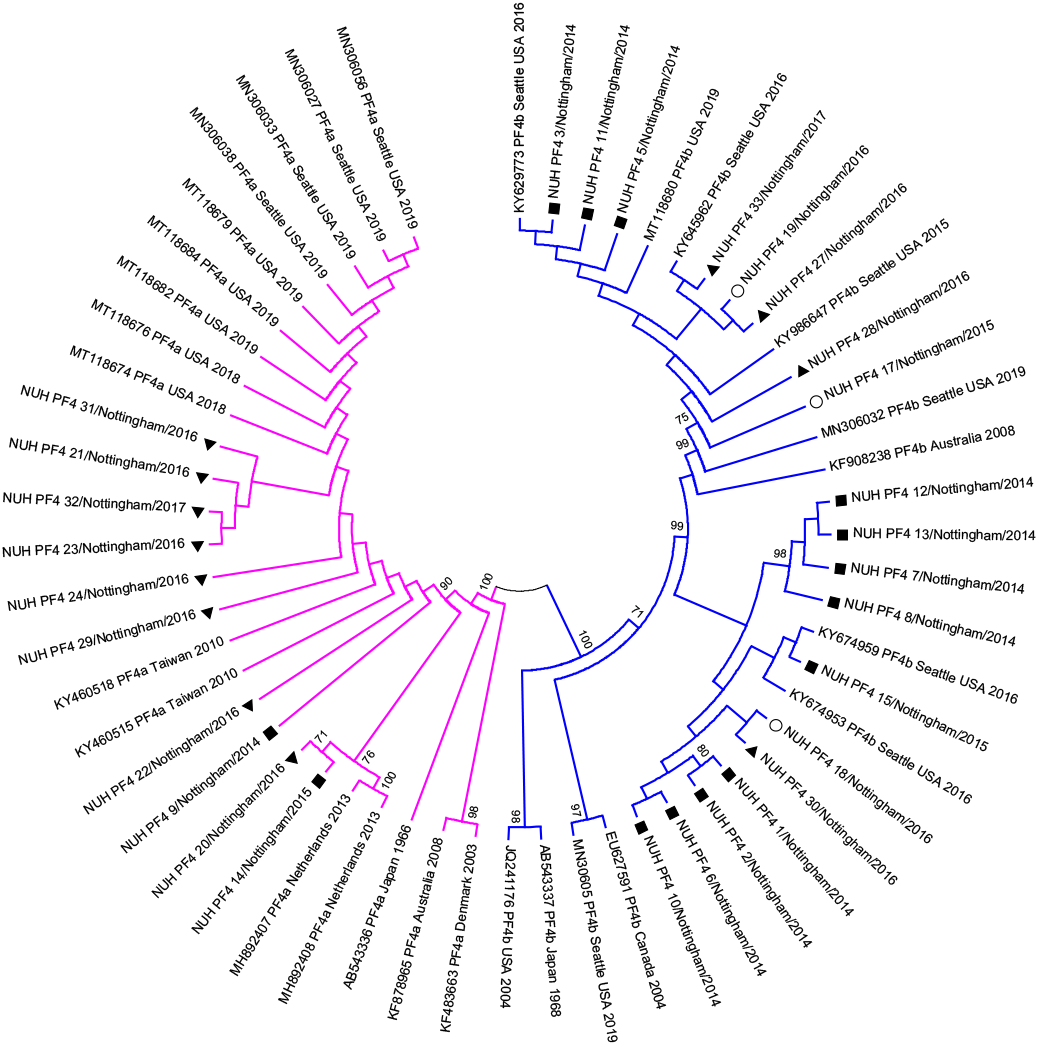
Phylogeny of HPIV4 study strains collected in Nottingham UK between 21^st^ November 2014 and 23^rd^ February 2017 compared to available reference sequences derived from GenBank in January 2020 under the taxonomic designation NCBI:txid 1979161. Sequences cover 793 nucleotides from 5310 to 6107 of HPIV4a reference strain M-25, GenBank Accession no. AB543336. The unrooted circular tree was constructed by the maximum likelihood method with 1000 bootstraps, displaying only tree topology; only bootstrap values with support greater than 70 are indicated. Filled squares and triangles represent study samples collected during the core HPIV4 2014/15 and 2016/17 epidemic seasons respectively, whilst open circle samples were collected in the autumn of 2015. HPIV4a (pink) and HPIV4b (blue) subtype designations are annotated by subtree branch colour. 2014 study sequences NUH_PF4_1 & 2, and 12 & 13 represent paired samples from the same patient.

### Clinical characteristics

Significant incidence of both co-infection and outpatient assessment was observed in the cohort, and HPIV infection could potentially be coincidental to a primary medical condition. Therefore, a focussed clinical audit was undertaken to compare features of hospitalised HPIV2 and HPIV4 mono-infected patients (56 and 75 individuals respectively) where routine diagnostic investigation could reasonably rule out mild and co-infection, to reduce confounding effects (Tables 1 & 2). However, in general, there were proportionally fewer hospital admissions for HPIV4-positive individuals (p<0.0001, data not shown).

Although almost twice as many HPIV4 mono-infected patients under the age of 10 were observed, the median ages were ultimately not statistically significant (p=0.0527, Table 1). Most strikingly, a fever was observed with more than 3-fold frequency in HPIV2 than HPIV4 patients (p<0.0001, Table 1) but no further significant differences were seen in variables assessed. For both HPIV2 and 4-infected and hospitalised patients, underlying medical conditions were very common as was immunocompromise (Table 2). Other than the aforementioned fever associated with HPIV2, shortness of breath and cough were the most commonly recorded symptoms, but intervention with nebulisers and supplementary oxygen was only required in a minority of instances; antibiotics were provided to approximately half of the cohort (Table 2).

**Table 2:** Investigations of hospitalised HPIV2 & HPIV4 mono-infected individuals

|  | **HPIV2** | **HPIV4** | **P Value** |
| --- | --- | --- | --- |
| **Characteristic** | **No. (%)** | **No. (%)** |  |
| **Radiological investigations** |  |  |  |
| Chest X-ray performed | 28 (50) | 43 (57.33) | 0.4791 |
| Radiological evidence of infection* | 15 (55.56) | 17 (38.64) | 0.2205 |
| **Haematological investigations** |  |  |  |
| Blood tested | 45 (80.36) | 53 (70.67) | 0.2284 |
|  | *low/normal/high* | *low/normal/high* |  |
| *Haemoglobin level\** | 26/18/1<br>(57.8/40.0/2.2) | 17/34/2<br>(32.1/64.2/3.8) | **0.0146** |
| *Platelet count\** | 14/27/4<br>(31.1/60/8.9) | 9/41/3<br>(17.0/77.4/5.7) | 0.309 |
| *Red Blood cell count\** | 24/20/1<br>(53.3/44.4/2.2) | 4/49/0<br>(7.6/49/0) | **<0.0001** |
| *White Blood cell count\** | 17/19/9<br>(37.8/42.2/20) | 1/52/0<br>(1.9/98.1/0.0) | 0.1293 |
| *Neutrophil count\** | 17/15/13<br>(37.8/33.3/28.9) | 7/25/21<br>(13.2/47.2/39.6) | **0.0224** |
| *Lymphocyte count\** | 25/19/1<br>(55.6/42.2/2.2) | 25/27/1<br>(47.2/50.9/1.9) | 0.461 |
| *Eosinophil count\** | 31/14/0<br>(68.9/31.1/0) | 22/27/4<br>(41.5/50.9/7.55) | **0.0028** |
| *Basophil count\** | 9/35/1<br>(20/77.8/2.2) | 12/41/0<br>(22.6/77.4/0) | 0.575 |
| *Monocytic cell count\** | 10/32/3<br>(22.2/71.1/6.7) | 2/45/6<br>(3.8/84.9/11.3) | **0.0136** |
| **Liver and kidney assessment** |  |  |  |
| Liver Function tested | 37 (66.1) | 33 (44.0) | **0.0139** |
|  | *normal/high* | *normal/high* |  |
| *AST level\** | 32/2 (94.4/5.6) | 31/1 (9.0/3.0) | >0.999 |
| *ALT level\** | 29/7 (80.6/19.4) | 25/8 (75.8/24.2) | 0.772 |
| *Bilirubin level\** | 33/3 (91.7/8.3) | 27/6 (81.8/18.2) | 0.294 |
| *Albumin level\** | 9/27 (25/75) | 5/28 (15.2/84.9) | 0.377 |
| CRP tested | 40 (71.4) | 43 (57.3) | 0.1039 |
|  | *normal/high* | *normal/high* |  |
| *CRP level\** | 14/26 (35.0/65.0) | 28/15<br>(65.1/34.9) | **0.0084** |
| Kidney Function tested | 44 (78.57) | 42 (56) | **0.0091** |
|  | *low/normal/high* | *low/normal/high* |  |
| *Sodium level\** | 8/35/1<br>(18.2/75.6/2.3) | 11/28/3<br>(26.2/66.7/7.1) | 0.765 |
| *Potassium level\** | 5/39/0<br>(11.4/88.5/0) | 6/31/5<br>(14.3/73.8/11.9) | 0.3279 |
| *Urea level\** | 11/26/7<br>(25.0/59.1/15.9) | 11/22/9<br>(26.2/52.4/21.4) | 0.7615 |
| *Creatinine level\** | 7/29/8<br>(15.9/65.9/18.2) | 4/30/8<br>(9.5/71.4/19.1) | 0.5464 |
| **Cerebrospinal fluid tested** | 2 (3.57) | 7 (9.33) | 0.2994 |

Patients identified between 1<sup>st</sup> September 2013 and 11<sup>th</sup> April 2017. \* Where data available for variable. Abbreviations: AST Aspartate Aminotransferase, ALT Alanine Transaminase, CRP C-reactive protein; p values <0.05 highlighted in bold

Patient blood was assessed more frequently in general than chest x-rays, with significantly more haematological aberrance outside normal parameters (i.e. both low and high levels, Supplementary Table 2) seen in HPIV2 but not HPIV4 patients. Specifically, more frequently observed low haemoglobin levels (p=0.0146) and low counts for red blood cells (p<0.0001), neutrophils (p=0.0224), eosinophils (p=0.0028) and monocytes (p=0.0136) were significant features of HPIV2 but not HPIV4 infection. Conversely, no differences were observed between HPIV2 and 4 for platelets, white blood cells, total lymphocytes and basophils (Table 2).

Liver and kidney function were assessed more frequently in HPIV2 patient care (p=0.0139 and 0.0091 respectively, Table 2). However, of the many parameters tested, a significant disparity was seen only in the more frequently abnormally high C-reactive protein levels of HPIV2 infection (p=0.0084, Table 2). In general, abnormally high albumin but not bilirubin levels were recorded whilst urea levels appeared marginally more frequently outside standard ranges than sodium, potassium and creatinine (Table 2).

## Discussion

In contrast to more prevalent pre-SARS-CoV-2 respiratory pathogens such as Influenza A and RSV, the Human Parainfluenza viruses are considerably under-studied, yet still present a significant burden to global healthcare [11, 24]. Previous key clinical studies have compared all four genetically distant types from different *Paramyxoviridae* sub-families together [9, 28], excluded certain age groups [9, 22] or overlooked HPIV4 entirely [22] and lacked complementary genetic investigation. Even reports addressing genetic analyses have been limited in both patient and sequence numbers and clinical detail [14-16]. The data presented here therefore represents to our knowledge the largest combined clinical and genetic study to date focussing exclusively on the epidemic human Orthorubulaviruses HPIV2 and 4.

Our retrospective observational period covered the principal epidemic autumn / winter period in 4 years, whereby we noted the biennial epidemic incidence previously attributed to HPIV2 [2, 3, 10]. A more complex pattern presented for HPIV4 was also consistent with other large-scale studies [3, 5, 11] alternating major and minor yearly epidemic seasons.

Many studies have previously observed a greater total incidence of diagnosed HPIV2 than HPIV 4 cases [28], including significant nationwide studies in the US [10] and the UK [3]. Notably the UK study deriving data from 158 laboratories between 1998 and 2013 observed twice as many positive HPIV2 than HPIV4 tests [3]. In contrast, we observed an almost 2-fold predominance of HPIV4 in general agreement with recent studies in China [6, 29], Vietnam [16] and the USA [9, 30]. This may be due to previous difficulty in culturing and related omission of HPIV4 from some diagnostic panels, increasing prevalence of HPIV4 or a combination of both factors [3, 10]. Notably Zhao and colleagues [3] detected a gradual increase of HPIV4 from 1998 through 2013, thus our contrasting results in the immediately subsequent period of 2013 to 2017 may actually indicate a further shift in the prevailing epidemiology of the human Orthorubulaviruses, in the UK at least. Continued uptake of HPIV4 in routine RT-PCR surveillance and national-level data aggregation [3, 10] will further elucidate the true incidence and seasonality of HPIV4 and its prevalence relative to HPIV2.

Although greater sequence coverage allows more in-depth resolution and interrogation of parainfluenza molecular epidemiology [14], even our relatively limited genetic investigation demonstrated contemporary co-circulation of genetically distinct subtypes of both HPIV2 and 4 in each of the four yearly epidemic seasons. This finding is consistent with previous investigation of HPIV2 & 4 in Vietnam [16] and Croatia [14]. Furthermore the predominance of Nottingham ‘V94-like’ HPIV2 sequences was also seen in Vietnam and Croatia, where a shift from ‘G3’[14] or ‘clade 1’[16] sequences analogous to our ‘V98-like’ designation between 2009 and 2014 to G1a / clade 2 / V94-like between 2014 and 2017 was observed [31]. This apparent pattern of genotypic replacement may be driven by population level immunity and susceptibility [31], however the clade under-represented by European sequences post 2014 was conversely over-represented by an unpublished cohort of reference sequences from Seattle, USA (NCBI Bioproject PRJNA338014).

HPIV4 genetic epidemiology is much less understood, with a considerable sequence archive paucity relative to even HPIV2, to such an extent that we were able to compare our 29 Fusion gene sequences to only 28 publicly available references. Although somewhat paradoxically, and in contrast to HPIV2, the apparent clades of HPIV4 have been accepted as subtypes A and B in the literature. This may have arisen by chance through early isolation of distinct HPIV4 strains in contrast to highly similar early HPIV2 strains [17-20], although HPIV4A & B are not considered to be distinct species [12]. Our data indicates the apparent increase in HPIV4 incidence in the UK [3] and possibly elsewhere involves multiple lineages of both HPIV4A and B, which we and others have demonstrated can cause clinical disease [32]. Even this most populous genetic study of HPIV4 to date is however under-powered to explore whether these subtypes and subtype lineages have different clinical properties, so like others [24] we would urge for increased sequencing allied to future clinical studies, alongside population level serological investigation. Fluctuating and currently changing climate conditions may also have a current and future role in the clinical and genetic epidemiology of parainfluenza and other respiratory infections [33, 34]. We found HPIV infections in all ages, with a more even distribution across age groups for HPIV2 in contrast to the more pronounced excess of HPIV4 in the under 9 and over 40 year old extremes of age, in general agreement with the epidemiological profile seen in the immediately preceding 15 year time period in the UK [3].

HPIV4 is classically described as a widespread but mild, self-limiting infection in contrast to HPIV2 with a strong etiological and epidemiological association with croup in infants (reviewed in [2]). Whilst we did see proportionately less hospitalisation with HPIV4-infected individuals, overall severity was comparable to HPIV2 in our mono-infected and hospitalised sub-cohort with a similar need for intervention with nebulisers and oxygen, alongside shortness of breath. Similarly Frost et al [9] noted more hypoxia in HPIV4 positive individuals compared to HPIV2 in children and generally similar severity between HPIV types. Fever was seen much more predominantly as a feature of HPIV2, but not HPIV4, infection, a trend previously noted by others, but without significance [9, 28] perhaps due to cohort limitations. C reactive protein has previously been noted as collectively elevated by HPIV infection [35], and mildly elevated during HPIV4 and not HPIV2 infection in children. In contrast, we found a significant elevation collectively in children and adults in HPIV2 and not HPIV4 infection.

Our findings of widespread antibiotic use for HPIV positive patients, even in the absence of non-viral co-infection are alarming but unfortunately not unusual [9, 22, 26]. This highlights the continued need for vigilant antibiotic stewardship particularly in the often-complex picture of HPIV infection with the frequent underlying medical conditions and immunocompromise observed.

A few limitations were apparent in our investigations. Clinical details were not recorded for the purpose of this study and thus recorded in a non-uniform and sometimes incomplete manner, reducing the availability of useable data and in turn statistical power. Even in these circumstances were able to achieve numbers of HPIV2 and 4 mono-infected individuals requiring hospital treatment equivalent to or exceeding the previous largest in-depth cohorts published [9, 16, 22, 28]. Collectively all these studies and ours fail to investigate the complete epidemiological burden of HPIV infections, with access only to clinical cases [31]. Extended community surveillance of asymptomatic or sub-clinical infection would greatly enhance knowledge of circulating HPIV strains, seasonal prevalence and associated pathogenesis, but will require considerable sampling effort [36]. Potentially related to limited pathogenesis in the community is the paucity of reference material with which to inform primer design and potentially sample degradation in storage, we, like others [16] struggled to amplify certain portions of the HPIV genomes. Furthermore, many of the archetypal [13] and also some more contemporary reference sequences [14, 15, 37] have been generated from cell cultured viruses, which may cause genomic changes and potentially affect chosen PCR priming sites. The additional primers and sequences described in this study, particularly for HPIV4, provide further tools for future PCR-based studies, whilst increasingly prevalent use of alternative deep sequencing strategies will further enhance knowledge of parainfluenza sequence diversity [38]

In summary, we found HPIV2 and 4 in the East Midlands of the UK between 2013 and 2017 to be caused by multiple co-circulating viral clades in adults and children. Both HPIV2 and 4 were frequently associated with hospitalised patients and occasionally severe disease. The general picture of respiratory disease in these individuals was distinguished by more frequent fever, abnormal haematology and elevated C reactive protein in HPIV2 positive individuals.

With the recent disturbance to typical transmission of endemic human respiratory viral infections caused by non-pharmaceutical intervention measures taken to control the SARS-CoV-2 pandemic, future orthorubulavirus epidemic patterns are uncertain and should be monitored carefully [39]. However the exceptional focus applied to the understanding and treatment of COVID-19 could yield advances in the management of patients with HPIV2 and 4 [40].

## Data Availability

All data produced in the present study are available upon reasonable request to the authors

## Acknowledgments

None to report

## Disclaimers

None to report

## Supplementary information

**Supplementary Table 1:**
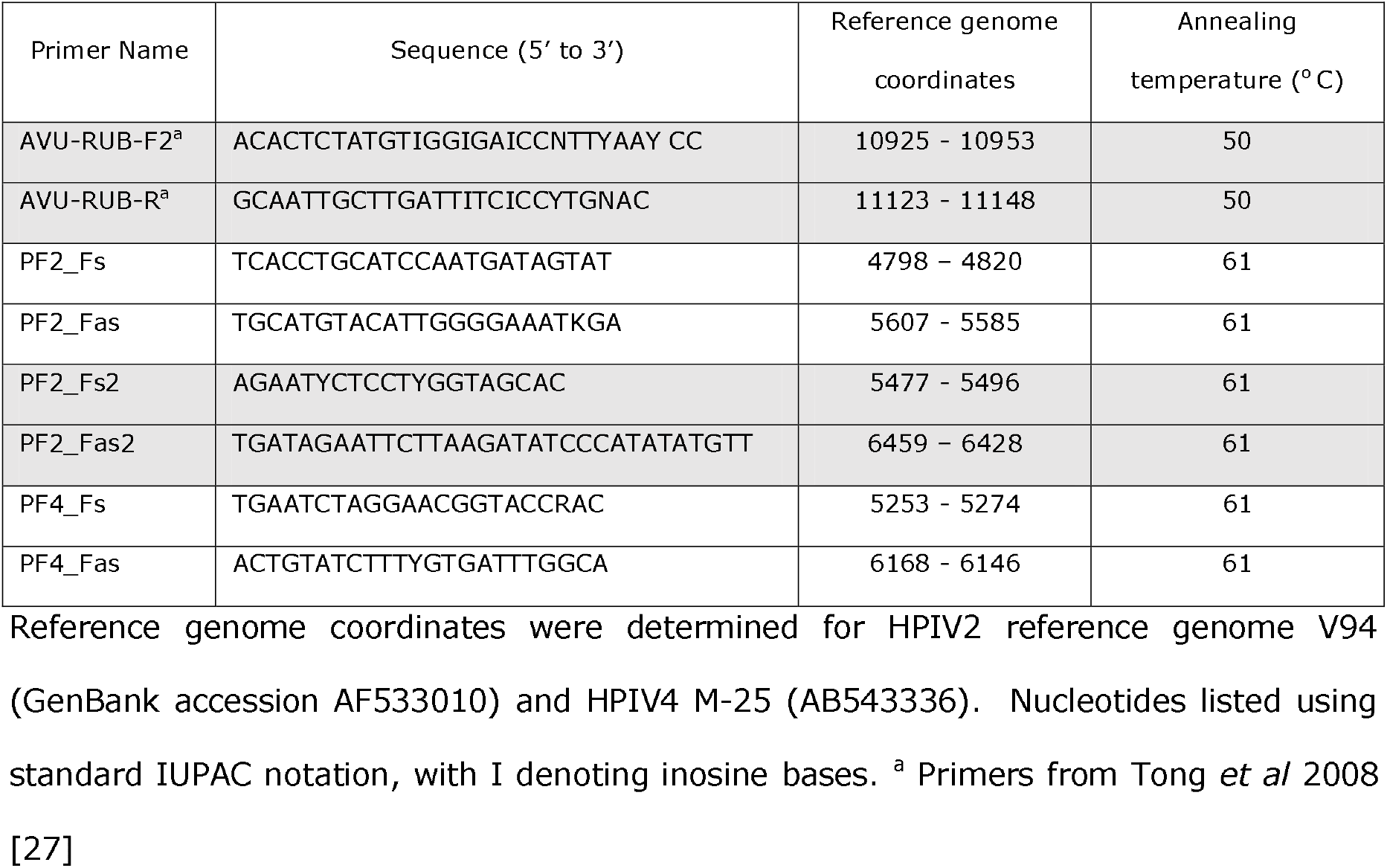
PCR Primers utilised in the study.

**Supplementary Table 2:**
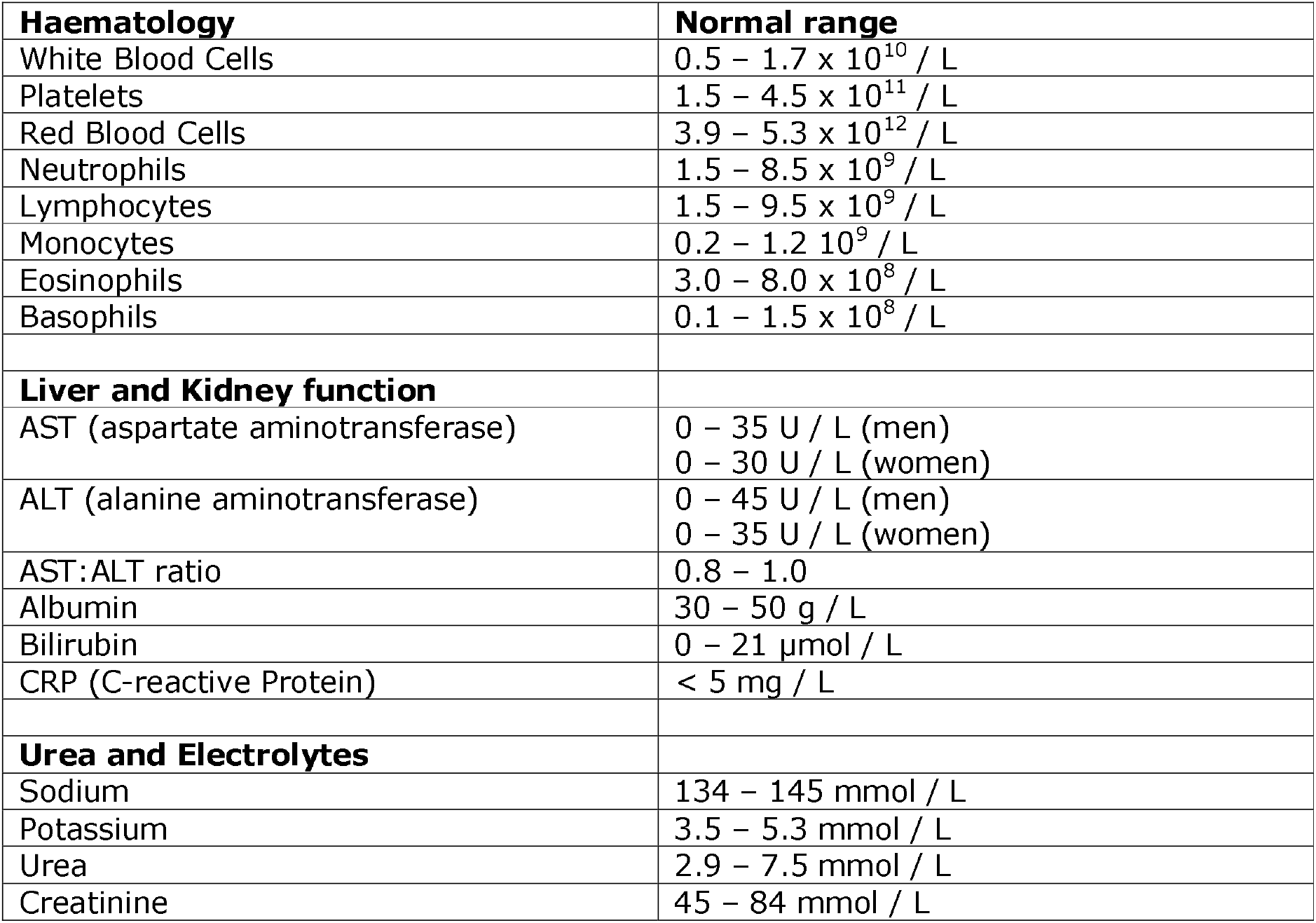
Ranges of normal values used for auditing of clinical features

